# The effect of reactive balance training on responses to novel unexpected balance perturbations: a feasibility study

**DOI:** 10.1101/2024.02.11.24302069

**Authors:** Andrew Huntley, Alison Schinkel-Ivy, Avril Mansfield

## Abstract

**Trial design:** Pilot study embedded within an assessor-blinded parallel randomized controlled trial.

**Objective:** To determine the feasibility of using unexpected and novel balance perturbations to assess the efficacy of reactive balance training.

**Methods:** <u>Participants</u>: Community-dwelling adults with chronic stroke who could walk independently without a gait aid for at least 10 m. <u>Interventions</u>: Reactive balance training, using manual and internal perturbations, or ‘traditional’ balance training (control group). Training took place for one hour per session, twice per week for six weeks. <u>Outcome</u>: Proportion of unexpected slips triggered as intended; **s**tate anxiety, perceptions of situations, and participants’ subjective responses to the unexpected slip perturbation; and spatiotemporal and kinematic features of unperturbed and perturbed walking (step length, width, and time, and mechanical stability) pre- and post-training. <u>Randomisation</u>: Blocked stratified randomization. <u>Blinding</u>: Assessors were blinded to group allocation.

**Results:** <u>Numbers randomised</u>: 28 participants were randomized (15 to reactive balance training, 13 to control). Of these, nine reactive balance training group participants and seven control participants were eligible and consented to additional data collection for the pilot study. <u>Numbers analysed</u>: 12 participants (six per group) completed the post-training unexpected slip data collection and were included in analysis of the pilot objective. <u>Outcome</u>: All unexpected slips triggered as intended.

Overall, participants did not report increased state anxiety or any concerns about the unexpected slip. Analysis of spatiotemporal and kinematic data suggested better stability following the unexpected slip for reactive balance trained participants than control participants; however, there were also between-group differences in spatiotemporal and kinematic features of walking pre- and post-training.

**Conclusions:** Unexpected slips are feasible in research. However, their value as outcomes in clinical trials may depend on ensuring the groups are balanced on prognostic factors.

**Trial registration:** ISRCTN05434601

**Funding:** Canadian Institutes of Health Research.

## INTRODUCTION

Reactive balance training (RBT) involves participants experiencing repeated loss of balance, with the goal of improving reactive balance control and preventing falls in daily life [1]. While there is evidence that RBT can prevent falls in daily life, study findings are varied [1]. The causes of falls are multi-factorial, and the occurrence of falls can be compounded by various factors beyond balance abilities (e.g., environmental and psycho-social factors). For example, an intervention that improves balance control may also improve balance confidence, leading to increased physical activity and/or risk taking, resulting in increased fall rates [2]. Therefore, rather than relying on fall rates alone to determine efficacy of RBT, researchers and clinicians should also examine the effect of RBT on reactive balance control.

Often, loss of balance during RBT is caused by external perturbations; perturbations can be provided manually by a therapist [3,4], or using a device such as a programmable treadmill [5,6], moving platform [7,8], or custom slip/trip walkway [9]. To determine generalizability of RBT, reactive balance control should be assessed using a different balance perturbation method than that used in training [10]. Participants who completed RBT using manual perturbations had fewer in-lab falls [4] and multi-step reactions [11] in response to moving platform balance perturbations than control participants. Likewise, participants who completed RBT using a programmable treadmill had greater improvements in the ‘reactive’ sub-score of the mini-Balance Evaluation Systems test [12] than control participants [6].

In daily life, balance reactions are typically executed in response to sudden and unexpected balance perturbations. In-laboratory or in-clinic reactive balance assessments typically do not mimic the unexpected nature of daily-life falls. While researchers or clinicians can introduce unpredictability into balance perturbations during assessment (e.g., in terms of perturbation timing or direction), participants are usually informed that the perturbation will occur. This expectation may increase anxiety or arousal, which can influence the nature of balance reactions [13]. Furthermore, people typically adapt their balance reactions quickly with repeated testing [14]. Therefore, characteristics of balance reactions observed during in-laboratory or in-clinic assessment, where participants expect the perturbations, and the same or similar perturbations are repeated multiple times, may not represent the reactions that participants execute due to an unexpected loss of balance in daily life [15]. Prior experience with the test perturbations pre-training may also influence performance on repeated testing of balance reactions post-training.

Reactions to truly unexpected balance perturbations can be evoked using deception protocols; that is, participants are explicitly told that they will not experience a balance perturbation, but the perturbation is provided [15-18]. The purpose of this pilot study embedded within a randomized controlled trial is to determine the feasibility of using unexpected and novel balance perturbations in clinical trials to assess the efficacy of RBT. Specifically, we aimed to determine if: the unexpected slip perturbations could be reliably provided; participants tolerated the unexpected perturbations (i.e., did not express any negative sentiments); and cross-sectional analysis of the outcomes would be useful in a future trial.

## METHODS

### Trial design

This is a pilot study embedded within a larger assessor-blinded parallel randomized controlled trial with 1:1 allocation ratio. The larger trial aimed to determine the effect of RBT on falls in daily life among people with chronic stroke. The study protocol and primary results for the larger trial have been reported elsewhere [3,10]. Eligible participants in the larger trial (see below) were invited to complete optional additional data collection using a motion platform [11]. The trial was approved by the University Health Network research ethics board (protocol number: 14-7428), and is reported according to the CONSORT extension for randomized pilot and feasibility trials [19].

### Participants

Participants were community-dwelling stroke survivors who could stand independently for ≥30 s without external support, and tolerate at least 10 lean-and-release balance perturbations [20]. Exclusion criteria for the larger study were: height/mass (limited by the safety harness system; >2.1 m and/or >150 kg); medical conditions that may have confounded the results (e.g., other neurological conditions) or posed an increased risk to participants (e.g., severe osteoporosis, or poorly controlled hypertension); communication or cognitive impairments that affected participants’ understanding of instructions; currently receiving physiotherapy or balance training; and/or completed RBT in formal rehabilitation less than one year previously. To be eligible for the additional motion platform component of the study, participants had to be able to walk independently for >10 m without a gait aid. Following baseline assessment, eligible participants were invited to undertake additional assessments on a motion platform prior to and after RBT. Participants provided written informed consent for the larger study and the additional assessment.

### Interventions

The interventions are described in detail elsewhere [3,21]. Briefly, all participants completed two one-hour training sessions per week for six weeks. Interventions were administered one-to-one by a physiotherapist (i.e., one physiotherapist per participant) in a research laboratory in an academic hospital.

The control group completed the Keep Moving with Stroke program [22] – a circuit-based exercise program for community-dwelling people with stroke. Each session included a 5-10 minute warm-up, 40 minutes of mobility and balance exercises (e.g., walking, sit-to-stand, heel raises, tap-ups or step-ups, reaching and weight shifting, and standing with reduced base of support), and a 5-10 minute cool-down with stretching.

RBT sessions included a 5-10 minute warm-up, voluntary tasks intended to induce internal balance perturbations, voluntary tasks combined with external balance perturbations, and a 5-10 minute cool-down. Participants wore a safety harness attached to an overhead support. Internal balance perturbations occurred when participants failed to control balance during voluntary movement. External balance perturbations were caused by a push or pull from the physiotherapist. Participants completed ∼60 balance perturbations per session, with the task difficulty set such that participants required an upper extremity response, external assistance (i.e., from the overhead harness or physiotherapist), or a multi-step response ∼50% of the time. Task difficulty progressed as participants improved with training.

### Outcomes

Data were collected pre- and post-training within FallsLab at the Toronto Rehabilitation Institute. FallsLab includes a 6-m long walkway mounted on a motion platform that can be programmed to translate in any direction [11]. Four force plates (AMTI, Watertown, Massachusetts, USA) were embedded in the centre of the walkway. Kinematic data were collected using a 13-camera motion capture system (Vicon MX40+, Vicon Motion Capture Systems Ltd., Oxford, UK) at 100Hz. Participants were asked to wear tight-fitting shorts and t-shirt and comfortable low-heel running shoes. Participants were outfitted with reflective markers on the acromion processes, lateral and medial humeral condyles, ulnar and radial styloid processes, greater trochanters, lateral and medial femoral epicondyles, lateral and medial malleoli, heels (most posterior aspect of the foot), 2^nd^ and 5^th^ metatarsal-phalangeal joints, and the tip of the big toe. Marker clusters were also placed on the arms, legs, and trunk, and seven markers were placed on the platform.

Participants first completed perturbations to stance and quiet standing balance trials that are reported elsewhere [23,24], and then performed three unperturbed walking trials. At the end of the post-training data collection session only, participants completed a fourth unexpected slip trial [16]. For all gait trials, including the slip trial, participants were explicitly told that the platform would not move to disturb their balance, although they were aware that the platform was capable of moving and had experienced moving platform balance perturbations earlier in the session. Participants were instructed to walk at their self-selected speed from one end of the walkway to the other. During the unexpected slip trial, forward platform movement was triggered when the vertical force on the front force plate under the less affected limb exceeded 50 N [25]. The perturbation waveform consisted of a square wave with 300ms acceleration followed by 300ms deceleration (peak acceleration: 1.5m/s^2^, peak velocity: 0.45m/s, total displacement: 0.135m) [16]. For all trials, participants wore a safety harness attached to an overhead track that followed them while moving around the platform.

Following the unexpected slip perturbation, the research team debriefed participants, informing them of the purpose of the deception (to provide a truly unexpected balance perturbation). Participants were invited to withdraw their data from the study if they were uncomfortable with the deception component of the study. They were also invited to report any concerns with the deception to the principal investigator and/or the institution’s research ethics board. After the debrief, participants completed the Endler Multi-dimensional Anxiety Scale (EMAS) – State and Perception [26]. The EMAS-Perception includes open-ended questions that ask participants to describe the situation they are in and to describe anything they felt was threatening about the situation or experience (if applicable).

Kinematic data were labelled using Vicon Nexus software (version 2.12.1, Vicon Motion Capture Systems Ltd., Oxford, UK), and analyzed using Visual 3D (version 2021.09.1, C-Motion, Germantown, USA). All kinematic data were low pass filtered at 6 Hz (body markers) or 10 Hz (floor markers) using a 2nd order dual-pass Butterworth filter. Whole-body centre of mass (COM) was calculated using an 11-segment model [27] (head-trunk, upper arms, forearms, thighs, shanks, and feet segments). COM and heel velocity were the derivative of COM and heel position, respectively, minus floor marker velocity. Margin of stability (MOS) was calculated using COM position and velocity, relative to the posterior edge of the base of support (heel marker), as described by Hof et al. [28] Heel contact time was the time when the anteroposterior velocity of the heel marker was <0.1 m/s. Heel contact events were visually inspected in Visual 3D and corrected, if necessary. For the slip trial, slip foot contact was the time when the less-affected limb contacted the force plate, thereby triggering the platform movement. Recovery heel contact was the subsequent heel contact time of the contralateral limb. We identified a corresponding less-affected heel contact in the unperturbed walking trials (i.e., the heel contact at a similar location on the moving platform), and subsequent contralateral limb contact time. Step time was the difference between the less-affected and contralateral limb contact times. Step length and width were the anteroposterior and medio-lateral distances, respectively, between the heel markers at contralateral limb contact (the step width sign was reversed for left-side affected participants to yield positive step widths when the limbs did not cross midline). Walking speed was the COM velocity at less-affected limb contact, and MOS was extracted at contralateral limb contact.

Global responses to the unexpected slip included whether >30% body weight was supported by the harness following the unexpected slip (i.e., a ‘fall’) [29], whether participants experienced a negative step length/negative overall gait velocity in response to the slip (i.e., a backward loss of balance), and whether participants displayed an upper-extremity response (e.g., sudden raising/swinging of arms) to help maintain balance in response to the slip perturbation (termed ‘arm response’).

### Sample size

We had no *a priori* sample size target for the pilot data collection. All eligible participants in the larger trial were invited to complete additional data collection for the pilot study.

### Randomization and blinding

Participants were assigned using blocked stratified randomization with allocation concealment to either the control or RBT group by the principal investigator, who was not involved in recruiting, assessments, or intervention administration. Variable block sizes of four, six, or eight were used. There were four strata from two stratification factors: site (two levels), and frequency of ‘failures’ during baseline reactive balance control assessment[30] (two levels). The random allocation sequence was computer generated and maintained in an electronic file by the principal investigator. Outcome assessors were blinded to group allocation.

### Analytical methods

We calculated the proportion of slip trials that were triggered as intended. EMAS-State scores were compared pre- and post-training using paired estimation plots [31] to determine if the unexpected slip trials increased anxiety among participants. EMAS-Perception scores were used to determine participants’ perception of the situation; scores >3 on each domain indicated that participants perceived that situation. Participant responses to EMAS-Perception open-ended questions were also examined for any negative sentiments. Between-group effects for kinematic and spatiotemporal gait data were calculated as the mean difference between groups and the corresponding 95% confidence interval, for pre- and post-training unperturbed walking trials and for the unexpected slip trials. Global responses were presented descriptively for each group.

## RESULTS

### Recruitment

Only eligible participants who were recruited into the larger trial from April 2015 onwards were invited to participate in data collection for this pilot trial. Sixteen participants completed additional data collection pre-training during this time. Two of these declined post-training data collection and two did not complete the unexpected slip trial (one due to technical issues unrelated to the slip perturbations, and for one participant we decided to not perform the unexpected slip perturbation as the participant strongly disliked the perturbations to stance). Therefore, the unexpected slip perturbation trial was attempted with 12 participants (six per group; Figure 1); all 12 participants are included in analysis of all outcomes. Participant characteristics are shown in Table 1. Data collection for the pilot trial ended when recruiting targets for the larger trial were met in August 2016 [3].

**Table 1:** Participant characteristics. Values presented are medians with ranges in parentheses for continuous or ordinal data.

|  | Control group | RBT group |
| --- | --- | --- |
| Age (years) | 63 (43-72) | 69 (51-78) |
| Sex (number) |  |  |
| Female | 2 | 1 |
| Male | 4 | 5 |
| Height (cm) | 169.2 (148.6-179.1) | 169.7 (162.5-178.0) |
| Mass (kg) | 74.5 (60.4-82.0) | 74.8 (69.7-87.9) |
| Time post-stroke (years) | 2.7 (0.5-14.4) | 2.4 (0.5-9) |
| More affected side (number) |  |  |
| Left | 4 | 5 |
| Right | 2 | 1 |
| NIH-SS stroke scale (score) | 4 (2-8) | 2 (1-7) |
| CMSA (score) |  |  |
| Leg | 5 (5-7) | 4.5 (3-6) |
| Foot | 4.5 (3-6) | 5.5 (4-7) |
| ABC (score) | 89.7 (34.4-96.3) | 90.9 (81.3-100) |
| BBS (score) | 52.5 (48-56) | 56 (51-56) |
| Mini-BEST (score) | 19.5 (14-23) | 22.5 (18-25) |
| TUG (s) | 10.9 (8.7-24.8) | 8.8 (5.6-22.9) |
| History of falls (number) | 2 | 2 |

ABC: Activities-specific Balance Confidence scale [32]; BBS: Berg Balance Scale [33]; CMSA: Chedoke-McMaster Stroke Assessment [34]; mini-BEST: mini Balance Evaluation Systems Test [12]; NIH-SS: National Institutes of Health Stroke Scale [35]; RBT: reactive balance training; TUG: Timed-up and Go [36]

**Figure 1:**
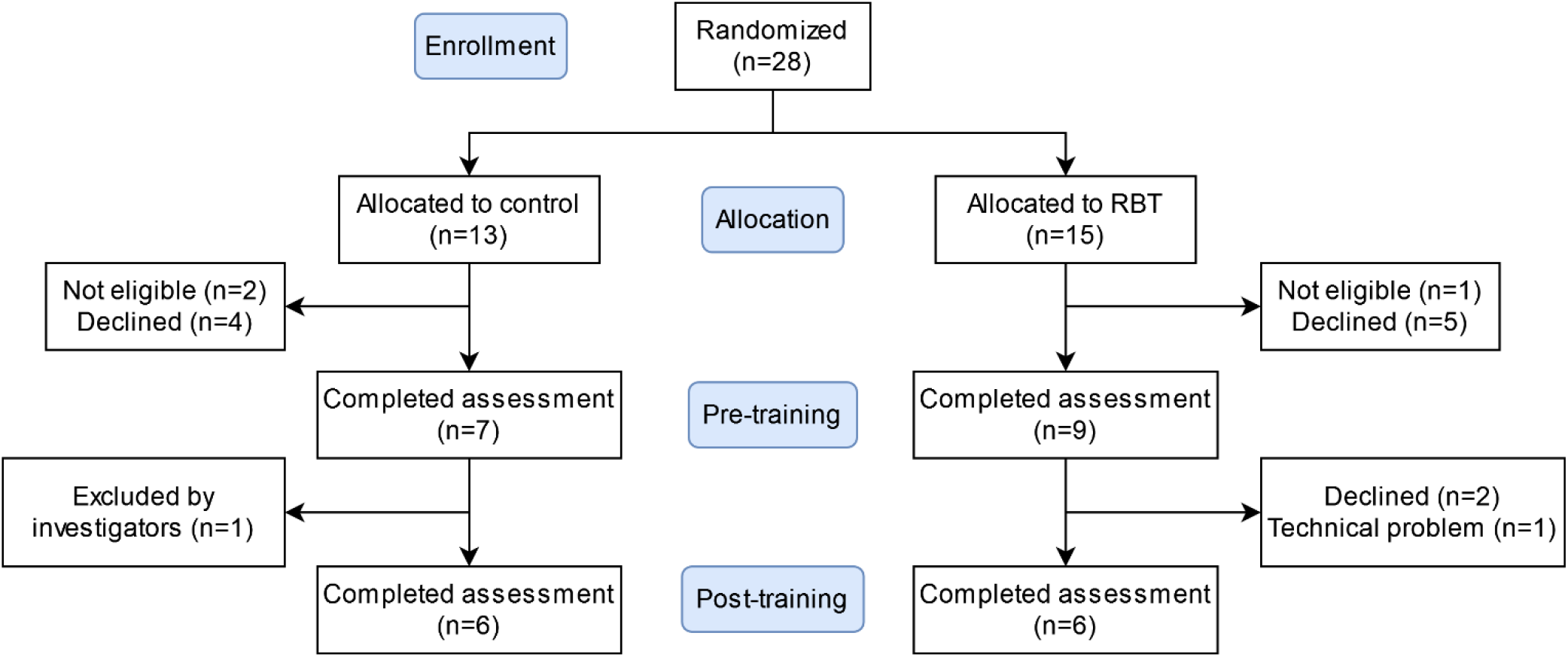
Participant flow diagram. Of the participants recruited into the larger trial between the dates when the pilot data collection was conducted, 16 were eligible and consented for the additional data collection. Two (RBT group) declined post-training testing. For one participant (RBT group), there was a technical problem unrelated to the unexpected slips that prevented us from collecting data as intended post-training. For one participant (control group), the research team decided not to attempt the unexpected slip perturbation as the participant disliked the perturbations to stance provided earlier in the session.

### Unexpected slip feasibility

The unexpected slip triggered successfully on less-affected foot contact for all 12 participants. On average, participants did not report increased state anxiety following the unexpected slip (post-training) compared to following unperturbed walking only (pre-training; Figure 2). The paired mean difference in EMAS-State scores between pre- and post-training was 0.17 (95% confidence interval: -2.5, 7.3; *p*=0.97). However, one participant in the control group increased their EMAS-State score from 32 pre-training to 55 post-training; this participant did not write anything in response to the open-ended questions to explain this increase in state anxiety. EMAS-Perception scores indicated that participants generally perceived that they were in a situation where they were being evaluated or were in a neutral situation, both pre- and post-training (Table 2). The participant who reported high EMAS-State scores reported perceiving that they were in a situation where they were in physical danger post-training (scored 4/5 for this item), although this participant also reported perceiving that they were in a neutral situation (scored 4/5 for this item). No participant reported perceiving that they were in a threatening situation, either pre- or post-training. Responses to the open-ended questions on the EMAS-Perception were generally positive both pre- and post-training (e.g., “interesting”, “excited”, “relaxed”, “happy”, “confident”, “fun”, “comfortable”). Post-training, one participant (RBT group) noted that they felt the harness “kicked in too early” and slowed them down. Another RBT participant noted post-training that they felt better and that they had improved. No participant commented on the unexpected slip perturbations during the debrief, nor did they report any concerns with the deception protocol to the principal investigator or research ethics board.

**Table 2:** Perceptions of situations. Values presented are the numbers of participants who perceived that situation (EMAS-Perception scores >3 for that item). Note that numbers do not add to 12 as some participants perceived that they were in more than one situation, and some participants did not perceive that they were in any of the situations presented.

|  | Pre-training | Post-training |
| --- | --- | --- |
| Situation where they are being evaluated | 7 | 4 |
| Situation where they are in physical danger | 0 | 1 |
| Novel, ambiguous or unfamiliar situation | 1 | 1 |
| Innocuous or neutral situation | 3 | 4 |
| Threatening situation | 0 | 0 |

**Figure 2:**
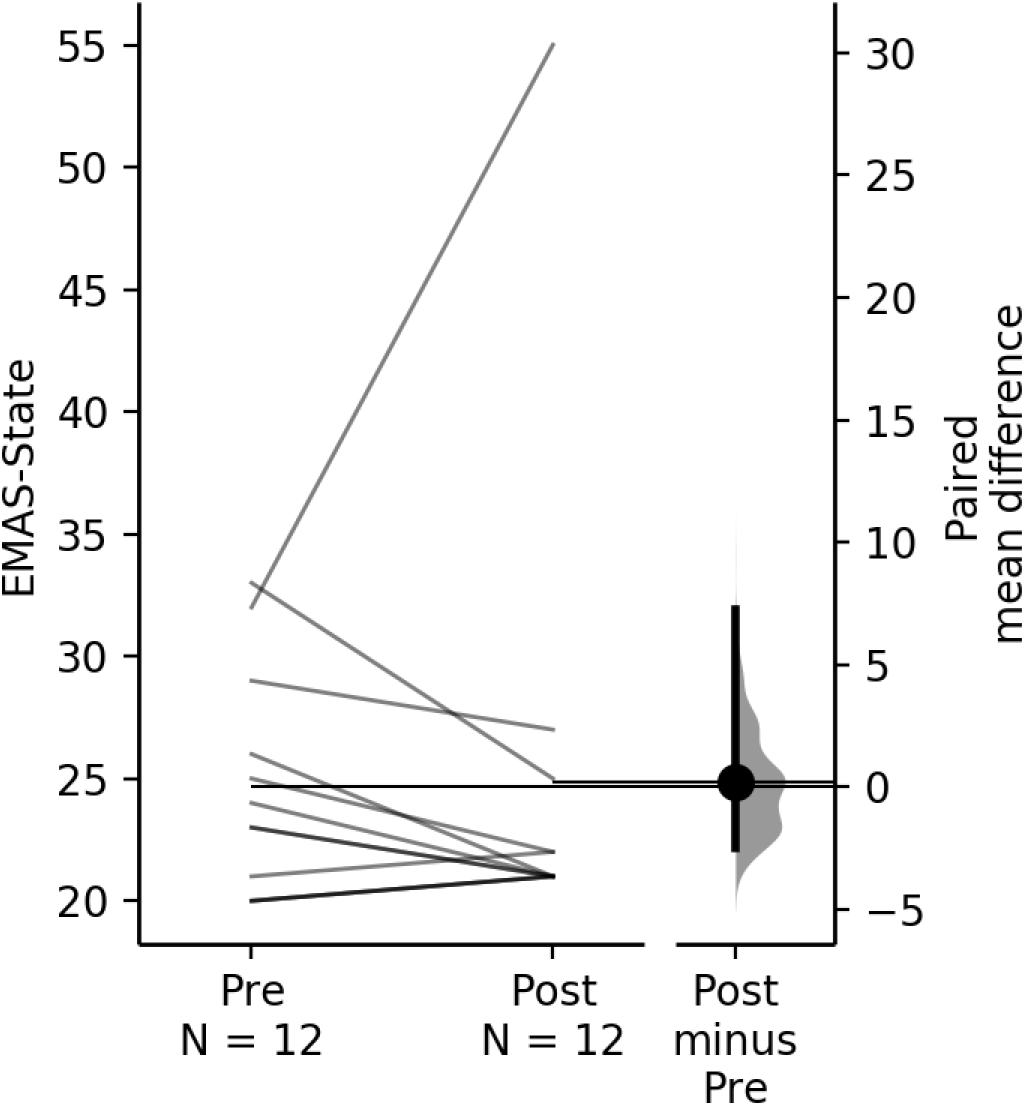
Endler-Multi-dimensional Anxiety Scale – State scores. The paired mean difference between pre- and post-training is shown in the above Gardner-Altman estimation plot [31]. Data for both sessions are plotted on the left axes as a slopegraph: each paired set of observations is connected by a line. The paired mean difference is plotted on a floating axis on the right as a bootstrap sampling distribution. The mean difference is depicted as a dot; the 95% confidence interval is indicated by the ends of the vertical error bar.

No participants from either group experienced a fall into the harness (>30% body weight) following the unexpected slip perturbation. Three participants in the control group (out of six) exhibited a backward loss of balance when responding to the unexpected slip perturbation, while only one participant in the RBT group (out of six) exhibited a backward loss of balance. Five participants in both groups exhibited an arm response to the slip perturbation. RBT participants appeared to have increased margin of stability and narrower step widths than control participants at recovery foot contact following the unexpected slip perturbation (Table 3). However, RBT participants also tended to have increased margin of stability and narrower steps than control participants at more-affected heel contact during unperturbed walking, both pre- and post-training.

**Table 3:** Spatiotemporal and kinematic variables pre- and post-training. Values presented are group means with standard deviations in parentheses, and between-group differences (RBT minus control) with 95% confidence intervals in brackets. Data are presented separately for unperturbed walking trials pre- and post-training, and unexpected slip trials (post-training).

|  | Trial | Control group | RBT group | Difference |
| --- | --- | --- | --- | --- |
| Walking speed (m/s) | Pre-training | 0.8 (0.3) | 1.0 (0.3) | 0.2 [-0.1, 0.6] |
|  | Post-training | 0.8 (0.3) | 1.0 (0.2) | 0.2 [-0.1, 0.6] |
|  | Slip | 0.8 (0.3) | 1.0 (0.2) | 0.2 [-0.1, 0.6] |
| Step time (s) | Pre-training | 0.74 (0.15) | 0.65 (0.08) | -0.09 [-0.25, 0.07] |
|  | Post-training | 0.69 (0.16) | 0.61 (0.07) | -0.09 [-0.24, 0.07] |
|  | Slip | 0.63 (0.19) | 0.60 (0.07) | -0.03 [-0.21, 0.16] |
| Step length (cm) | Pre-training | 44.2 (12.8) | 54.7 (14.3) | 10.6 [-6.8, 27.9] |
|  | Post-training | 47.2 (11.1) | 56.5 (12.3) | 9.2 [-5.8, 24.2] |
|  | Slip | 34.9 (35.8) | 45.8 (22.6) | 10.9 [-27.6, 49.4] |
| Step width (cm) | Pre-training | 12.2 (2.8) | 8.8 (4.4) | -3.5 [-8.2, 1.2] |
|  | Post-training | 13.8 (5.2) | 8.0 (4.4) | -5.8 [-12.0, 0.4] |
|  | Slip | 17.1 (5.9) | 10.3 (4.6) | -6.8 [-13.6, 0] |
| Margin of stability (cm) | Pre-training | 52.1 (18.0) | 73.1 (13.0) | 21.0 [0.9, 41.2] |
|  | Post-training | 60.0 (15.6) | 75.0 (13.1) | 15.0 [-3.6, 33.5] |
|  | Slip | 57.9 (12.9) | 74.1 (10.5) | 16.1 [0.1, 31.2] |

RBT: reactive balance training

## DISCUSSION

This pilot study aimed to determine the feasibility of unexpected slip perturbations as an outcome in clinical trials of balance training interventions. Among this sample of people with chronic stroke, the slip perturbation was feasible. The slips triggered as intended for all participants, and it appeared that participants did not have a negative perception of the deception protocol, which was required to provide an unexpected slip. Similar unexpected slips with deception have been used in studies that included healthy young [16,18] and older adults [15,17], with no reported concerns expressed by participants. While researchers can use a mix of perturbed and unperturbed walking trials to provide unpredictable perturbations, participants may alter features of their walking in anticipation of the perturbation [37]. Unexpected perturbations with deception (i.e., participants being informed that there will be no perturbation) allow researchers to provide in-laboratory balance perturbations that are more similar to the completely unexpected loss of balance that participants experience in their daily lives.

Despite the feasibility of the unexpected slip protocol, the data may have limited value in clinical trials. Between-group comparison of outcomes at the post-training time point alone would have led us to conclude that RBT led to improved stability (i.e., higher MOS and better global responses) in response to the unexpected slip perturbations than the control intervention, demonstrating generalizability of RBT. However, there were also apparent between-group differences in features of unperturbed walking, including higher MOS for the RBT group compared to the control group, both pre- and post-training. Therefore, it is unclear how much of the between-group difference is due to training effects versus pre-existing individual differences in characteristics of walking. Future trials using a post-intervention unexpected balance perturbation should use an allocation method that balances the groups on features of unperturbed walking (e.g., minimization [38] or stratified allocation [39]) to facilitate between-group comparison in outcomes post-intervention only.

To participate in the additional data collection, participants had to be able to walk at least a short distance without a gait aid. Therefore, participants in our study scored highly on tests of functional balance and mobility and balance confidence. The results of this study might not apply to participants with worse balance and mobility function; that is, the deception protocol used in this study may not be feasible or tolerable for people with more severe balance and mobility problems. The slips provided by our moving platform may not resemble real-world slip perturbations, as both feet move forward on heel contact, whereas slips typically involve just the front limb slipping forward. However, initial balance responses to this type of moving platform ‘slip’ are similar to slips that only perturb the front limb [16].

In conclusion, unexpected novel balance perturbations are feasible to include as outcomes in clinical trials. Researchers who wish to use unexpected balance perturbations in post-training only should carefully consider how to balance the groups on important prognostic factors to help increase confidence that any between-group differences observed are due to the intervention rather than differences between the groups at baseline.

## Data Availability

All data produced in the present work are contained in the manuscript

## Notes

**Funding:** This study was supported by the Canadian Institutes of Health Research (MOP 133577). The authors also acknowledge the support of the Toronto Rehabilitation Institute; equipment and space have been funded with grants from the Canada Foundation for Innovation, Ontario Innovation Trust, and the Ministry of Research and Innovation. These funding sources had no role in the design of this study and did not have any role during its execution, analyses, interpretation of the data, or decision to submit results.

### Competing Interest Statement

The authors have declared no competing interest.

### Clinical Trial

ISRCTN05434601

### Clinical Protocols

https://bmcneurol.biomedcentral.com/articles/10.1186/s12883-015-0347-8

### Funding Statement

This study was supported by the Canadian Institutes of Health Research (MOP 133577). The authors also acknowledge the support of the Toronto Rehabilitation Institute; equipment and space have been funded with grants from the Canada Foundation for Innovation, Ontario Innovation Trust, and the Ministry of Research and Innovation. These funding sources had no role in the design of this study and did not have any role during its execution, analyses, interpretation of the data, or decision to submit results.

### Author Declarations

The trial was approved by the University Health Network research ethics board (protocol number: 14-7428).

